# Understanding older people experiencing homelessness with complex health and social circumstances (PHECHS): Qualitative study

**DOI:** 10.64898/2026.03.18.26347969

**Authors:** Mabhala Mzwandile

## Abstract

**Objective:** The study aimed to understand the perspectives of professionals from multi-agencies working with individuals experiencing homelessness with complex health and social circumstances (PHECHS), specifically focusing on how they define and contextualise the concept of “complex needs”.

**Method:** sixteen qualitative interviews with multi-agencies working with people experiencing homelessness were analysed using Heidegger’s interpretive phenomenological analysis (IPA), theories of socioeconomic determinants and international and national policy analysis were utilised to analyse data collected by MM on the multi-agency approach to individuals experiencing homelessness with complex health and social needs (PHECHS).

**Findings:** The analysis of a multi-agency approach aimed at supporting PHECHS revealed that complex needs arise gradually during childhood and can continue into adulthood. A range of factors contribute to both homelessness and these complex needs. Deconstructing the social and economic factors that underpin this continuum is essential to effectively addressing these challenges. This study conceptualises the complex needs of PHECHS into two key themes: deconstructing the PHECHS and attritional approach to PHECHS.

**Conclusion:** Homelessness is a grave human rights violation, depriving people of essentials like housing, food, health, education, and social participation. Governments have a moral and legal duty to end homelessness. Real progress demands comprehensive, sustained, and rights-based strategies that tackle root causes—poverty, trauma, and social exclusion. Homelessness also stems from gaps in vital life skills: job seeking, financial management, accessing services, and self-care. These barriers make it even harder to secure stable housing. Lasting reductions in homelessness result from a strong legislative framework, national guidelines, and sustained financial investment.

**Strengths and limitations of this study:**

1. Employing qualitative methods and analysing data through the lens of socioeconomic determinants of health inequalities enabled the development of a model that clarifies the structural causes of homelessness and highlights key opportunities for preventive policy interventions.
2. Examining the data through the lens of socioeconomic health determinants reveals how systemic and structural factors, such as housing policy and service access, drive complex needs beyond individual circumstances.
3. The study was conducted in an affluent, demographically homogenous city, resulting in the underrepresentation of individuals from ethnic minority backgrounds, women, and young people.
4. Future research should investigate the experiences of people experiencing homelessness using an asset-based perspective, leveraging frameworks that emphasise resourcefulness to promote their meaningful engagement and inclusion in society.

## 1. Introduction

Homelessness is intertwined with complex health and social care needs [1]. People experiencing homelessness with complex needs include individuals with a history of trauma, mental health disorders, those facing disadvantages due to age or life transitions (such as care leavers), people fleeing abuse and violence, and those who are multiply disadvantaged by poverty and inadequate housing [2, 3]. This group also includes individuals who are marginalised, at high risk, or hard to reach—such as those involved in substance misuse, offending, or at risk of social exclusion. Additionally, people with a dual diagnosis (such as co-occurring mental health disorders and substance misuse) or other combinations of medical and mental health disabilities are included [4, 5]. The literature demonstrates that these population groups experience disproportionately higher rates of homelessness [1, 6, 7]. Importantly, it is often the combination of two or more of these disadvantages or complex needs that precipitates the most severe form of homelessness: rough sleeping [6, 7]. For example, the literature indicates a significant rise in homelessness among individuals with mental health disorders, with an 83% increase reported in 2024 [1]. Therefore, the data highlight the need for services addressing homelessness to encompass the multifaceted health and social needs of this population [1].

The literature provides substantial evidence that complex needs arise from a dynamic interplay of interrelated structural (poverty, social, economic, and political) and intermediate (material circumstances, psychosocial, behavioural and biological) factors.[6–11]. As demonstrated by Harland et al. [8] and Mabhala et al. [6], the intersection of adverse life experiences and mental health disorders is a critical foundation for the development of complex needs. For example, both Mabhala et al [6] and Harland et al. [6]found that adverse childhood experiences—such as abuse, bereavement, and family instability—when compounded by mental health disorders, poverty, and housing instability, often lead to adulthood characterised by multiple and intersecting disadvantages. To conceptualise these multifaceted challenges, Keene [9] identifies five broad domains of complex needs: psychological issues, learning disabilities, social problems, involvement in crime, and substance misuse. Building on these insights, Rankin and Regan [10] argue for a holistic approach to care, emphasising that effective interventions must address the individual as a whole rather than focusing on isolated conditions.

Further supporting this perspective, Macías Balda [12] highlights that individuals with complex needs typically face three or more interrelated challenges, such as mental health illness, substance misuse, physical disabilities, and homelessness. Across the literature, poverty emerges as a pervasive factor that exacerbates these complexities [5–7, 11]. For instance, Padgett et al. [11] emphasise that among those affected by poverty and homelessness, individuals with serious mental health illnesses encounter additional barriers, including stigma and social exclusion. These findings collectively signify the importance of addressing poverty as a central strategy in tackling the multifaceted nature of complex needs.

Despite the well-documented link between homelessness and complex needs, the literature lacks a comprehensive and universally accepted definition of ‘complex needs,’ revealing a notable gap in its understanding. Existing definitions, such as those provided by the National Institute for Health and Care Excellence [13], typically focus on individuals who require extensive support across multiple aspects of daily life, often due to illness, disability, or other life circumstances. These definitions fail to capture the full complexity and intersectionality of the challenges individuals face, particularly the dynamic interplay among psychological, physical, and social factors. Furthermore, there is limited attention to how structural determinants—such as systemic inequality, housing policy, or access to services—shape the experience and manifestation of complex needs [2, 3, 6, 14, 15]. This highlights the need for a more nuanced and holistic conceptualisation that recognises both individual and structural contributors to complex needs.

This study aims to systematically investigate the perspectives of multi-agencies engaged with individuals experiencing homelessness and complex health and social circumstances (PHECHS). A primary focus is placed on elucidating how these professionals define and contextualise the concept of “complex needs.” To achieve this aim, a comprehensive literature review was conducted to analyse the current understanding of homelessness among people with complex needs in the UK. Theoretical frameworks related to socioeconomic determinants were applied to critically analyse the evolution of UK policy responses to homelessness. This approach offered nuanced insights into the shift in policy priorities that have characterised governmental strategies over time. Empirical insights were gathered from professionals across multiple agencies supporting individuals experiencing homelessness and complex needs. These perspectives were systematically analysed to clarify the definitions of complex needs and to evaluate proposed strategies for effective intervention. To the best of our knowledge, this is the first study to address homelessness through an approach that combines policy analysis, socioeconomic determinants, and human rights frameworks with empirical insights from multiple agencies. This research is original in its integration of theory, policy, and practice, providing a clearer, multidimensional understanding of complex needs in homelessness. Its significance lies in the potential to guide more integrated policy solutions and targeted interventions, offering valuable contributions to both scholarship and real-world strategies for addressing homelessness.

### 2. Method

The study used Heidegger’s interpretative phenomenological analysis, theories on the socioeconomic determinants of health, and a rights-based approach to synthesis the perspectives of people working with individuals experiencing homelessness. These methodological approaches enable a nuanced exploration of how structural and systemic factors shape the lived experiences of those facing homelessness and complex health and social challenges. Applying the socioeconomic determinants framework to PHECHS is crucial for understanding the interplay among social, economic, and environmental factors that influence health outcomes and overall well-being [77, 78, 85]. Furthermore, a human rights-based perspective asserts that homelessness infringes on several basic rights: social security, adequate food and housing, employment, living standards, health care, education, and participation in cultural life [16]. Recognising these interconnected issues is essential for creating sustainable measures to tackle homelessness.

### 2.1 Setting and recruitment

This study was undertaken as part of the Multiagency Approach to Rough Sleeping (MARS), a collaborative initiative involving representatives from a range of organisations (Table 1 shows the composition of the MARS). MARS holds fortnightly meetings to discuss cases of individuals experiencing rough sleeping in the city, particularly those with complex needs. During one such meeting, the Principal Investigator formally presented the research proposal and invited all representatives to participate, providing contact details for those interested in volunteering. Individuals who contacted the research team and expressed interest received an information sheet and consent form. Sixteen participants ultimately provided informed consent to take part in the study. The characteristics of these participants are summarised in Table 1 below. The study received ethical clearance from the University of Chester, Health, Medicine and Society Subcommittee.

**Table 1:** Representatives and types of agencies represented in the multiagency approach to rough sleeping in local authorities in Northwest England.

**\* Pseudonyms have been used for anonymity**

### 2.2 Data collection

Sixteen semi-structured interviews were conducted with representatives of MARS by multiple researchers. To ensure methodological consistency, an interview guide—developed specifically for this study and not previously utilised. All interviews were audio-recorded and subsequently transcribed using Microsoft Teams. The duration of interviews ranged from 25 to 55 minutes.

### 2.3 Data analysis

The three core principles of Heidegger’s IPA—dasein (the perceived reality of being-in-the-world), idiographic (the study of lived experience), and hermeneutics (the interpretation of experience)—provide a rigorous framework for data analysis[17–20]. “Dasein,” as articulated by Heidegger, refers to being-in-the-world and highlights the necessity of attending to individuals’ lived experiences and self-awareness within their unique contexts [17–20]. This ontological stance asserts that reality and existence are subjectively perceived, varying across backgrounds and perspectives [17–20]. The idiographic approach focuses on the specific challenges and narratives of each participant, rather than producing generalised conclusions. [17–20]. Hermeneutics, as a qualitative approach, interprets participants’ narratives to uncover deeper meanings, acknowledging the researcher’s interpretive frame as both necessary and inevitable.[17–20] (87, 88). Building on these principles, data were organised using Benner’s thematic analysis [20], which operationalises dasein, idiographic, and hermeneutic approaches through two phases: data organisation and interrogation, and the interpretive and narrative stage.

**Figure 1.**
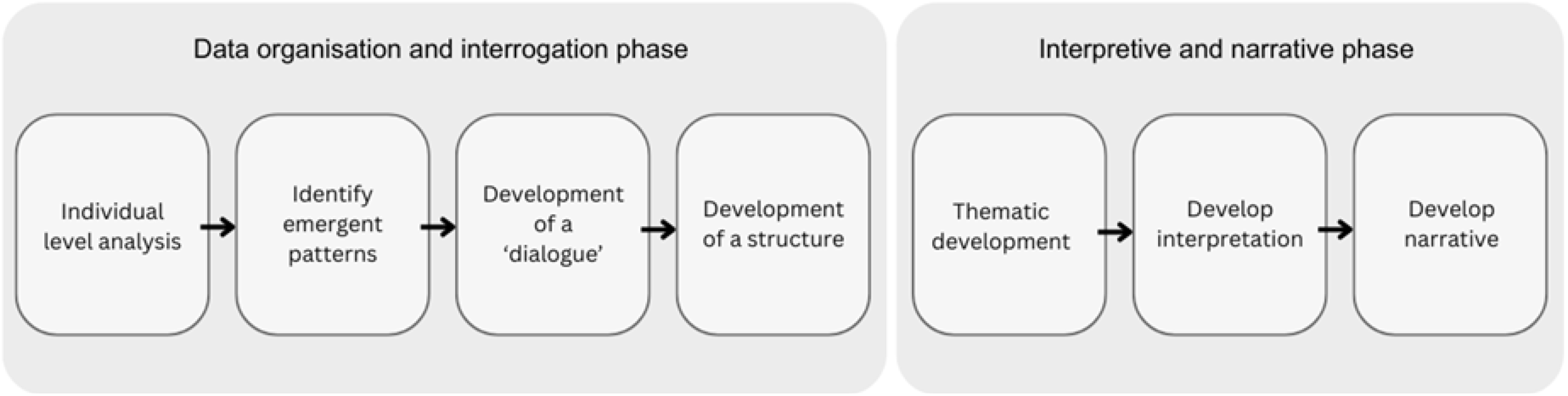
illustrates the two phases of the interpretive qualitative data analytic process used to analyse the data collected from March to July 2019 in Northwest England. The data used to populate this figure is extracted from Larkin and Thompson.

As demonstrated in Figure 1, the interpretive and interrogation phase began with a line-by-line analysis of individual participants’ experiences within a specific context.

We identified the emerging patterns of meaning that participants ascribed to their experiences. These experiences were then coded using concepts that captured the patterns observed. Following this, we explored the relationships among the different themes, posing questions about the significance of participants’ concerns within this context. Next, we developed a structural framework to illustrate the relationships between the themes. The second phase involved clustering themes with similar meanings into overarching themes and subthemes, along with providing theoretical explanations for these groupings [18].

## 3. Findings

This study identifies two key themes in the complex needs of PHECHS: deconstructing PHECHS and the attritional approach to PHECHS.

### 3.1 Deconstructing the PHECHS: continuum of Adverse Circumstances

There are three related subthemes that illustrate how adverse health, economic, and social circumstances impact individuals from childhood through adulthood: childhood trauma, adult experiences, and emerging descriptions of PHECHS. Figure 2: Provides a visual representation of the paths to becoming an individual experiencing homelessness with complex needs.

**Figure 2:**
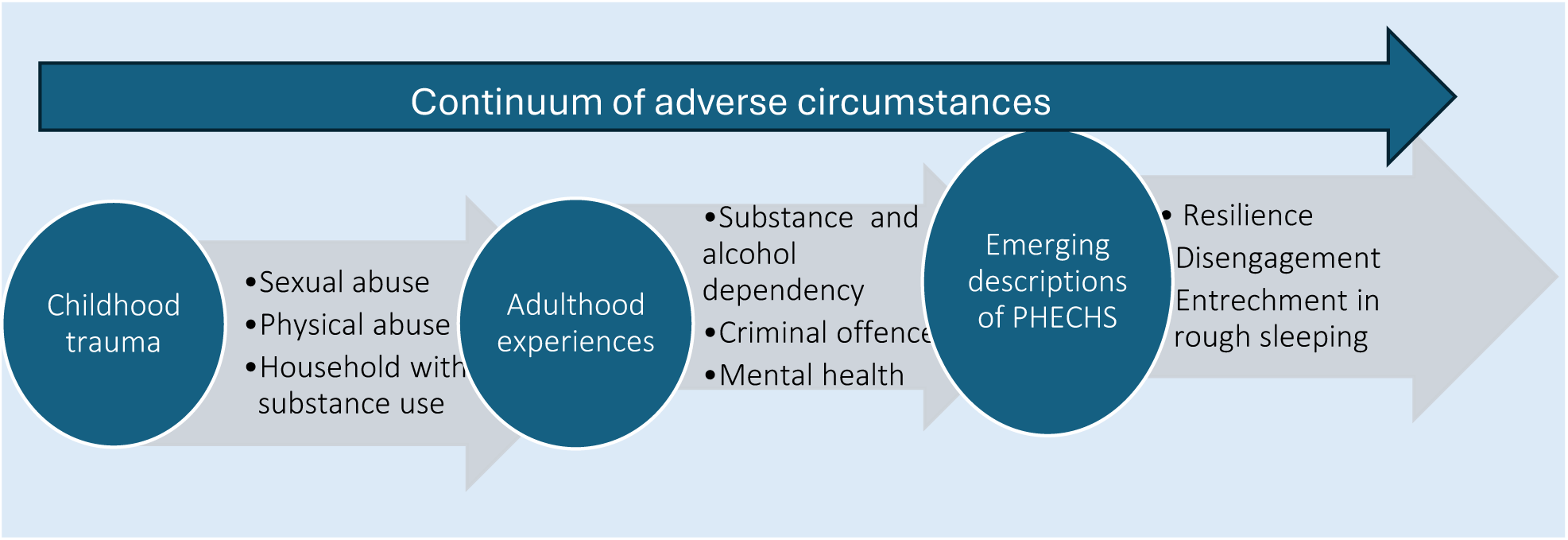
shows the paths to becoming an individual experiencing homelessness with complex needs.

**\* Pseudonyms have been used for anonymity**

The analysis revealed that PHECHS are defined as those facing multiple adversities simultaneously, impacting various aspects of their lives—including physical and mental health, social and economic well-being—over an extended period. These adversities often stem from childhood trauma and continue into adulthood; the complexity of these adversities necessitates a collaborative, multidisciplinary approach involving various professionals and sectors working together for a sustained period to address them effectively.

> *Complex needs mean many different things to different people. But in terms of homelessness, for most people, they’ve got two or more of the following: they would have sort of substance misuse issues, physical or mental health issues. They might have suffered significant traumatic events in their lives that have put them in their current position. [Silas]*

Participants in this study identified five key criteria that distinguish individuals with complex needs from the broader population experiencing homelessness. These criteria include:

1. Multiple adversities occurring concurrently that impact various facets of an individual’s life.
2. An extended duration of rough sleeping (entrenchment)
3. A history of traumatic experiences
4. Mental health disorders
5. Interaction with the criminal justice system

In this study, all participants identified individuals experiencing homelessness with complex needs as those facing a confluence of challenges that require a multifaceted approach for effective intervention. Such individuals typically grapple with various issues, including physical and mental health disorders, substance abuse, housing instability, difficulties related to social security or employment, as well as entanglements with the criminal justice system. The following excerpts provide clear examples of how participants typically describe complex needs.

> *Complex needs are, well, from my perspective, people with multiple significant needs, usually in lots of different domains, such as mental health needs, physical health needs, perhaps, and substance use needs. I would say they are individuals who have a variety of needs. It’s not just one issue they’re facing; there are multiple challenges occurring at the same time [Cleo]*

> *My role tends to be more with the women; I guess from my experience working with females there tends to be a lot of abuse history, whether that’s sexual abuse, emotional abuse, physical abuse, or domestic violence in their history and usually, some substance as well. [Cleo]*

In contrast to many individuals encountering homelessness who often utilise temporary shelters, PHECHS frequently endure prolonged periods of unsheltered living.

> *The fact that they are rough sleeping is a key difference between those with complex needs and others homeless groups. But another key difference is that it’s somebody with a long history of rough sleeping. [Torin]*

Participants explained that individuals experiencing homelessness typically sleep on the streets for periods ranging from 40 weeks to as long as 20 years.

> *So, examples would be where we’ve had people who have slept rough for over 40 weeks, but in the main, it’s people that we are trying to get in there early to try and get them back into mainstream services. [Silas]*

> *As soon as they’re accommodated, they come off it. Very quickly, often within weeks, they’re back on the streets. They’ve been in it for 20 years; you’re not going to get them out within six months. That’s the challenge of dealing with really entrenched, complex cases. [Cecil]*

In this study, all participants consistently identified a history of trauma as a pervasive factor among individuals experiencing homelessness with complex needs.

#### 3.1.1 Childhood trauma

In this study, all participants observed that PHECHS have experienced significant childhood trauma. The most common types of childhood trauma identified among PHECHS include sexual abuse, physical abuse and being raised in environments where important adults abuse substances or are involved in criminal activities.

> *You have childhood abuse, sexual abuse, physical, exposed to domestic violence. So, I’d say that in nearly all the cases they would be put into a trauma category to some extent or others. [Cecil]*

> *I would say all of them do have a history of trauma, some are more complex than others, and that can be trauma from either physical or psychological abuse from childhood into adulthood. It is not something that has just happened in one incident. It is usually multiple traumas that have happened in their lives from childhood until adulthood. [Alisha]*

Participants observed that childhood trauma experienced by PHECHS tends to persist into adulthood. Many individuals who endured childhood trauma tend to exhibit more propensity for poor decision-making.

> *I would like to provide an example of an individual I assist. This person has a history of abuse; they were abused as a child and have also experienced sexual and physical abuse as an adult. So, because of that, they have very limited trust in anybody. Trying to get that person engaged with drug and alcohol services isn’t easy, and because of that trust issue. [Cleo]*

Participants noted that most PHECHS grew up in households where drug use was prevalent. They believe that such an upbringing may have impacted their realities in adulthood.

> *I deal with an awful lot of people who have perhaps experienced trauma at an early age, and who have come from dysfunctional families, or where drugs or alcohol have played a part and subsequent deaths in the various kin have meant that they have. They have run away, moved away from their housing, or lost their original housing. [Catalina]*

> *They did not grow up living with their family, possibly because their parents were addicted to drugs and alcohol, which led the parents to distance themselves. [Lily]*

Participants observed that the health-harming behaviours exhibited by PHECHS in adulthood—such as drug and alcohol abuse, criminal activities, and antisocial behaviour—are rooted in childhood trauma.

> *Generally speaking, individuals who have experienced significant trauma in their lives, particularly in childhood, often resort to coping mechanisms such as alcohol and drugs. These coping strategies are usually the root cause of their challenges. [Diana]*

> *If you were an outsider observing them, you could see that they have a drug problem; it is obvious. However, the real challenge lies in understanding the reasons behind their drug and alcohol issues in the first place. Unlocking those reasons is the key to helping them. [Cleo]*

#### 3.1.2 Adult experiences

Participants identified several common reasons for PHECHS to lose their tenancies. These reasons include eviction from their homes or being asked to leave by family and friends. Factors contributing to this loss of support often involve drug use, alcohol dependency, involvement in criminal activities, mental health disorders and relationship breakdowns. Additionally, some individuals feel compelled to flee abusive situations, such as domestic violence.

> *Several factors come together, leading to the loss of their homes, and usually include drugs, alcohol, and offending behaviour. [Silas]*

> *The majority, most of all, either have issues with alcohol or drugs, and some of them do have underlying mental health issues [Mavin]*

They observed that the assessment of individuals facing substance use issues for accommodation can be particularly challenging, as many individuals are often under the influence of these substances during the evaluation process.

> *It can be difficult to get an assessment when someone is under the influence of drugs or alcohol. If they are often under the influence when agencies see them, it becomes hard to complete any social care or mental health assessments [Torin]*

Participants discovered that within their residences, PHECHS often engage in antisocial behaviours and exhibit aggressive conduct, which could lead to eviction due to the influence of drugs and alcohol.

> *So, a typical situation might be somebody who is under the influence of drugs or alcohol engage in antisocial behaviour [Torin]*

> *We’ve seen some customers go from a point where they’ve been actively involved in antisocial behaviour, drinking, taking drugs, shouting, and not engaging with service provisions to the levels where antisocial behaviours were completely reduced. [Silas]*

Moreover, many individuals decline to participate in drug and alcohol treatment services when offered these opportunities.

> *I think the real big problem around that is in terms of their use of drugs and alcohol, and the vast majority of them struggle to engage with sort of mainstream services. [Silas]*

> *Trying to get that person engaged with drug and alcohol services isn’t easy, and because of that trust issue [Cleo]*

> *Some of them will accept the accommodation, but then won’t want to accept their drug or alcohol misuse. [Cloe]*

All participants noted that among PHECHS, alcohol and substance use act as maladaptive coping mechanisms stemming from unresolved traumas faced during their formative years.

> *it’s people who have suffered a lot of traumas in sort of the background or their lives or maybe even in childhood, which tends to lead to them using alcohol and drugs as coping mechanisms to deal with things [Diana]*

The study highlighted that PHECHS are overrepresented in the criminal justice system. Some people get in contact with social services and services for people experiencing homelessness via the criminal justice system.

> *You know that again, this group of people overrepresented in terms of the criminal justice system is another point of engagement for some [Sam]. They often draw the attention of the police [Cloe]*.

> *Typically, they may have an offending history or antisocial behaviour [Torin]*

In certain rare situations, social services may refer PHECHS to the criminal justice system. This often happens when multi-agency teams identify serious criminal activity or antisocial behaviour.

> *So, you know, as the last chance scenario of somebody’s behaviours where they were causing significant criminality or significant antisocial behaviours, then the police would step in. However, the police should be considered a last resort rather than the first option. [Silas]*

> *In this field, the police officer needs to be a social worker, a health worker, and a support worker. [Silas]*

It was recognised that although referring individuals to the criminal justice system is sometimes necessary as a final option, relying solely on this approach may not effectively resolve the issues faced by PHECHS.

> *I want to return to the criminal justice issue, as it presents a complex dilemma. Our main priority is to provide support. Sanctions cannot be addressed in isolation; they must be implemented alongside support measures. While sanctions may not prevent this behaviour, our goal is to always consider support as the primary option rather than turning to sanctions as the first approach. [Silas]*

Participants observed that PHECHS almost always have underlying mental health disorders. These mental health challenges are significant factors that contribute to their difficulty in securing and maintaining stable housing.

> *They are unable to retain accommodation because of other elements of need that they have, and I think it is most usually a combination of mental ill health, probably likely to be substance and alcohol misuse or child. [Alice]*

> *The majority, most of all, either have issues with alcohol or drugs, and some of them do have underlying mental health issues. [Mavin]*

The experience of rough sleeping itself can inflict significant emotional and mental damage, which can further exacerbate existing mental health issues and lead to a cycle of instability.

> *Individuals who experience rough sleeping often suffer from mental and emotional damage as a result of their circumstances and how they ended up in this situation. Understandably, rough sleeping would take a toll on their emotional and mental well-being, leaving them in a difficult place. [River]*

The intertwined nature of mental health issues and substance use makes it difficult to diagnose behaviours accurately. Participants expressed concerns that individuals could be mislabelled with personality disorders rather than receiving proper mental health assessments.

> *I think there is a lot about personality disorders. We say it is not a mental illness. He does not have a mental illness; it is a personality…. There is a push for personality disorder labels. [Cecil]*

Mental health professionals play a vital role in reaching out to those in need, providing support without requiring stringent diagnostic criteria. This accessibility is critical for engaging individuals who have been disconnected from services.

> *My particular role is mental health; if we have got somebody who is homeless and they want to talk about their mental health, I will do it. There are no specific criteria or a diagnosis to refer; it can be from anyone. [Mavin]*

> *In the early days, there was a prevailing mentality surrounding mental health that often led people to believe that individuals should be detained under the Mental Health Act. Many assumed this was the best solution when someone was struggling. However, we have been able to challenge these misconceptions. We have worked to clarify that detaining someone in a psychiatric ward is not the answer and should be considered a last resort, if at all. It is essential to approach mental health with understanding and compassion rather than assumptions. [Diana]*

Improved communication between medical services and mental health professionals is crucial, especially in emergencies. Collaborations facilitate better identification and support for individuals in crisis, ensuring they receive the necessary care and assistance to help stabilise their situations.

> *The medical practice communicates with the hospital and the A&E department regarding mental health cases. One case involved a presentation about an individual’s mental health, highlighting the benefits of a multiagency programme. It has established connections with the Community Mental Health Team, which is vital when assessing individuals whose behaviour raises concerns from the police and the community. [Silas]*

> *In several instances, a multiagency programme has facilitated hospital admissions for individuals whose behaviour warranted intervention [Silas]*

#### 3.1.3 Emerging descriptions of PHECHS

Participants observed that people experiencing chronic homelessness (PHECHS) tend to disengage from services, express distrust toward professionals, and remain entrenched in rough sleeping. Over time, they develop resilience as a response to these challenges. This resilience, while a testament to human adaptability, often masks the systemic issues that necessitate more effective interventions and support frameworks.

Living on the streets requires a significant degree of functionality, resourcefulness, and endurance, as it is a 24/7 challenge. Those with complex needs exhibit adaptability to adverse conditions and often build supportive communities among themselves.

> *Despite complex needs, usually the people we work with are very resourceful. They’re very streetwise, and they’ve learned lots of ways to cope and live on the streets because it’s a very harsh and difficult environment to live in. So, they’ve adapted, you know, and have that resilience to cope and live out there because it is a tough world and being on the streets. [Cleo]*

> *They are resourceful and resilient, but sometimes, there can be anger and mistrust of the kind of services and professionals. [Cleo]*

PHECHS face substantial obstacles in accessing services considered standard within their societies. These obstacles can seriously hinder their engagement with, and reintegration into, the wider community. A significant challenge is the frequent mismatch between when professional support is offered and when individuals are actually ready to participate in such services.

> *Some people are not in a place where they want to engage with mental health at all. They just aren’t ready or want to. So, you get a lot of non-engagement and decline any engagement with services*.

> *Feel that’s a great need, but the person themselves doesn’t see it that way and would rather not engage. [Cleo]*

> *…. someone who probably does not engage or is refusing to engage. So, it takes extra services to get that person off the streets and then engage with services, which is the challenge. They may work with one person, but they might not want to work with another. [River]*

Another significant barrier is the negative past interactions that PHECHS have had with professional and social institutions. These past experiences not only shape their perceptions but also amplify their feelings of marginalisation by society and its institutions.

> *And they’ve trusted so many people in their past and have been let down. [Alisha]*

> *A lot of people, especially the that I work with, have had traumas associated with professionals they previously dealt with. Some have had a child removed. Perhaps they’ve just had negative encounters with professionals. They’ve usually had negative experiences with professionals previously. They want to reluctant to engage with professionals, certainly with a professional who they might view as having a potential problem person or part of their trauma history is difficult for them. [Cleo]*

In this study, nearly all participants used the term “entrenched” to describe the significant challenges faced by individuals experiencing persistent homelessness. The term refers to those who have experienced prolonged periods of homelessness, characterised by repeated cycles of rough sleeping and multiple unsuccessful attempts to secure stable housing. This group may have lived on the streets or in temporary accommodations for extended durations, leading to a deep embedding in their circumstances.

Entrenched homeless individuals often face significant barriers to housing due to a history of failed tenancies, which diminishes their viable housing options. They often exist on the fringes of society, feeling marginalised and disconnected from community resources and mainstream assistance programs.

> *I think the homeless population aren’t the same. Some are a little bit more entrenched. Maybe I’m a little bit more on the fringes of society. [Cecil]*

This subgroup typically exhibits resistance to traditional intervention strategies, complicating efforts to provide adequate support.

Participant describes a typical entrenched client with a history of numerous unsuccessful tenancies and attempts to secure stable accommodations, leaving them with few viable housing options.

> *Typically, it might be a customer who has dealt with housing options or homeless services several times previously, tried different options, maybe provided temporary accommodation, and, in some cases, even found more settled accommodation, and then the customer comes back again. [Torin]*

They often return to the support systems multiple times. Despite securing placements in rehabilitation or temporary housing, many face ongoing setbacks that hinder sustainable progress, resulting in a cyclical pattern of instability within the system.

> *Some people come around and go through the process multiple times. They would secure placements such as alcohol and drug treatment or rehabilitation, or we would get them into temporary accommodation. Unfortunately, quite a few of them hit a bump in the road or stopped halfway through. Unfortunately, we see that quite regularly with some of our more entrenched rough sleepers. [Cleo]*

They developed distinct communities; to enter them, one must earn their trust.

> *For many clients who are entrenched in this world, taking a little step into it can be pretty tricky at times, because they have trust issues — not easy straight away. [Alisha]*

Many entrenched homeless individuals are reluctant to engage with traditional intervention strategies, necessitating innovative, flexible approaches to reach and support them effectively.

> *I would say the rough sleeping and the entrenchment would probably be why people are considered on multiagency instead of the traditional homeless… people who are rough sleeping either refuse an offer or continue to rough sleep. [Torin]*

### 3.2 Attritional approach to PHECHS

A comprehensive, multilayered approach is thus essential in tackling homelessness. This approach must not only provide immediate housing solutions but also confront the underlying causes, including domestic violence, substance dependence, and both physical and mental health challenges. Furthermore, it must address fundamental determinants of homelessness, such as pervasive poverty, child poverty in particular, educational deficits, poor employment opportunities, and the enduring impact of adverse childhood experiences.

The study revealed that to ensure housing stability, service providers must adopt a comprehensive approach that extends beyond mere housing provision. They must assist individuals in developing essential daily living skills, such as impression management, cooking, cleaning, and home management. Equally important is the provision of hands-on support in navigating health and social services, including assisting with social security benefit claims and accompanying them to healthcare appointments. Furthermore, improving employment prospects is a key component that involves helping individuals with literacy and numeracy skills, writing curriculum vitae, job applications, and connecting them to employment services. All these elements are vital for ensuring long-term housing stability.

> *They want me to go with them to appointments and micromanage them. I’ll be outside on my bike. I’ll give them a wake-up call. I’ll take them there. You know, it depends on which direction they come from. [Lily]*

Participant emphasises that this basic support is vital for PHECHS as they lack the basic skill required to function in mainstream society.

> *Support is vital because some people cannot function in the mainstream. If somebody has been a long-term rough sleeper who is not engaged with any form of service to achieve success, they, after talking to the outreach staff on a very simplistic level, start to access the drug and alcohol service or see a doctor. For us, that would be a success. [Silas]*

Participants observed that addressing PHECHS requires sustained flexibility in support settings, which is critical for effective intervention. Instead of enforcing strict attendance policies that can lead to case closures and abandonment, a more successful approach prioritises continuous, adaptable assistance. This flexibility is essential, as rigid policies often force clients back onto the streets. By ensuring that support is consistently available and tailored to individual needs, a more compassionate and effective system can be created that prevents abandonment and promotes long-term stability.

> *We need to adopt a flexible approach …, because if I ask any of the clients to leave, their next option is often the streets. [Lily]*

> *Many organisations would have a three-strike rule, say, I have three strikes, and you’re out. For example, if you try to access healthcare and a doctor gives you three appointments, you fail to turn up for those three appointments, and they could close your case. We don’t do that in multiagency. We keep going, and we don’t give up. [Silas]*

Participants indicated that the PHECHS endeavours to ascertain potential agents’ commitment before engaging them. They want to ensure the agents will remain dedicated and do not abandon them during difficult times.

> *I will contact them by telephone and arrange a meeting; they’ll reschedule it at the last minute. I will be there again the following week and the week after that. And then eventually, when they know that I want to work and help them, they’ll start to engage. Their lives on the street are chaotic. [Alisha]*

Addressing PHECHS is characterised as an attritional approach. Service providers must continue their efforts, despite setbacks, to connect with individuals.

> *What we do on multiagency is very attritional; we don’t give up on people. However, they are very difficult client-based clients to work with, and an attritional approach is really important. If you took a traditional approach, like a set, an appointment failed to attend, or stopping your service, we wouldn’t be working with any of them. [Silas]*

> *An attritional approach is vital here; the process takes a long time. It’s not instantaneous. [Silas]*

“We never discharge them” illustrates the need for unwavering commitment to consistent support and long-term engagement with PHECHS, ensuring no one is left behind.

> *We never discharge them. We continually try to work with them and find new ways to work together and generate new ideas. They are used to being told no to and used to being disregarded. The fact that we continually keep working with them is a good example [Pearl]*

Participant illustrates the attritional nature of working with PHECHS.

> *It could be six months before they speak to and engage with anyone. This is where it doesn’t tick the funding boxes that you’re looking at in the long term [Pearl]*

Again, illustrating the attritional nature of working with PHECHS

> *The complex needs require very slow, long work. It takes an awfully long time to build rapport and trust with someone who has not had trust since probably the day they were born [Lily]*

Building trust through persistent outreach is not just critical; it is crucial. Individuals may take time to engage with services, and service providers need to demonstrate a sincere commitment to helping them.

Recognising that some people will require help indefinitely. Participants observed that certain aspects of the extensive support systems in place for PHECHS might require sustained support indefinitely. This is especially pertinent for those experiencing persistent mental health disorders, learning disabilities, and individuals who are particularly at risk of adult exploitation.

> *So, I think we, as a local authority, must appreciate that the customers needing support may need it indefinitely. So you’ve got to be realistic. They may also have learning disabilities. While they may be capable of living independently, their thought processes may not be as developed as others’, leading them to require more support [Lily]*

Participants observed that taking a trauma-informed approach allows the providers to explore the underlying reasons for behaviours associated with individuals experiencing homelessness. Rather than labelling behaviours like aggression as negative, this perspective encourages understanding the traumatic experiences that may contribute to such behaviours.

> *I am trauma-informed. I believe that these people should be understood, rather than saying he is being aggressive, we’re not seeing him again. We need to understand why he is being aggressive. What is behind that behaviour? What does that tell us about him? So that trauma-informed and trauma recovery as well, I think that’s quite embedded into my thinking [Cecil]*

A trauma-informed approach recognises that many individuals have experienced significant trauma, leading to difficulties in trusting others. Taking the time to build relationships through informal interactions, such as sharing a cup of tea, helps create a safe environment where individuals feel comfortable discussing their challenges. This gradual approach is essential in fostering openness and encouraging engagement with support services.

## 4. Discussion-Conceptual framework for PHECHS

This study set out to systematically investigate the perspectives of multi-agencies engaged with PHECHS, with a primary focus on elucidating how these professionals define and contextualise the concept of “complex needs.” Figure 3 graphically synthesises the two principal themes that emerged from this analysis, providing a nuanced illustration of the intricate interplay between individual and structural determinants of PHECHCS. It also indicates the key entry points for intervention at both the individual and structural levels to effectively address not only the immediate manifestations but also the underlying root causes of PHECHCS.

**Figure 3.**
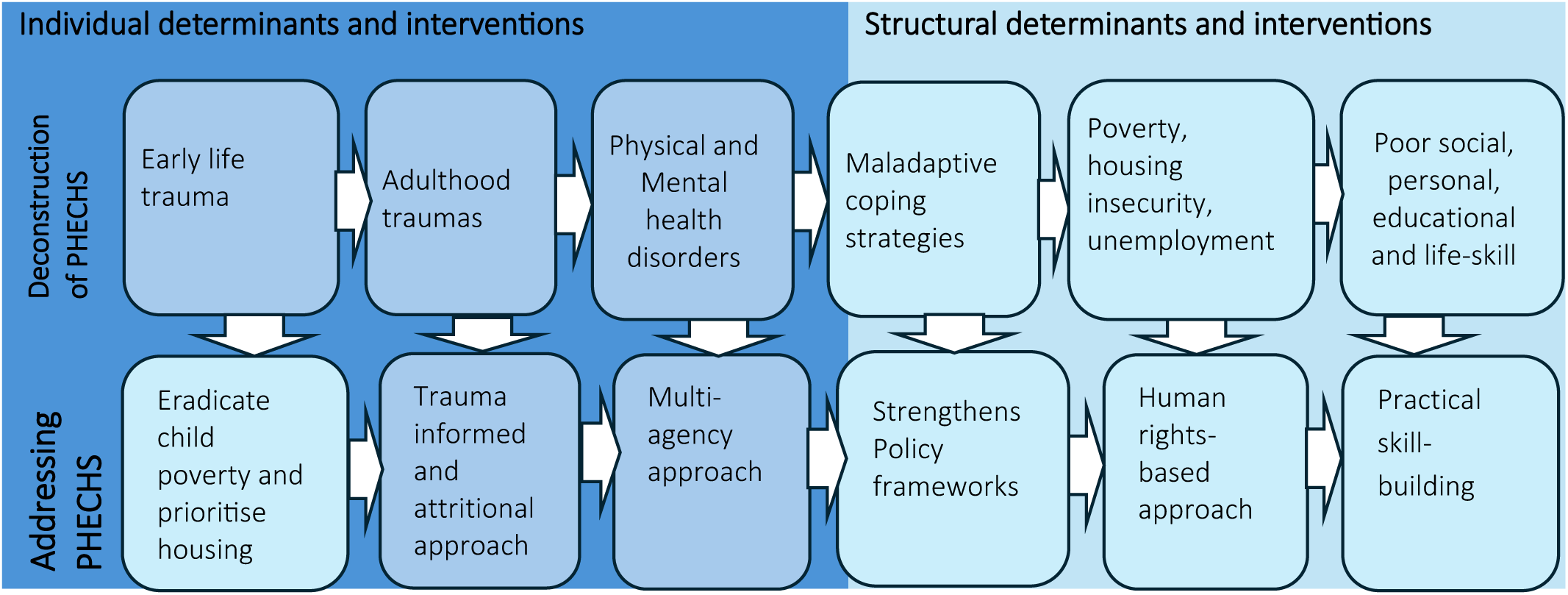
presents the conceptual Framework for explaining the Occurrence, Defining Attributes, and Measures to Address Homelessness with Complex Needs (PHECHS)

Figure 3 presents the conceptual Framework for explaining the Occurrence, Defining Attributes, and Measures to Address Homelessness with Complex Needs (PHECHS)

The analysis revealed that complex needs in the context of PHECHS are characterised by multiple adversities occurring simultaneously, affecting various aspects of their lives—including physical and mental health, social and economic well-being—over an extended period. Several researchers have defined complex needs in their professional contexts over the years, including Keane (2001), Rosengard (2007), and Balda (2016).

Though they differed in specificity and detail, they shared the common understanding that complex needs cannot be fully addressed by a single discipline[2, 9, 12]. The principal distinction between the present study and prior conceptualisations lies in its focus on the concurrent manifestation of multiple needs sustained over a prolonged duration.

A systematic analysis of multi-agency support for PHECHS identified two principal themes: (1) the deconstruction of PHECHS and (2) an attritional approach to addressing PHECHS. Regarding the first theme, participants in this study articulated five salient criteria distinguishing individuals with complex needs from the broader homeless population.

1. Multiple adversities occurring concurrently that impact various facets of an individual’s life.
2. An extended duration of rough sleeping
3. A history of traumatic experiences
4. Mental health disorders
5. Interaction with the criminal justice system

This finding is consistent with previous research: Keane delineated five defining criteria[9], Rosengard enumerated nine[2], and Balda [12] characterised complex needs as the co-occurrence of three or more interrelated issues, including mental illness, substance use, physical disability, and homelessness. Across these frameworks, mental health disorders, substance use, homelessness and poverty consistently emerge as core elements in the conceptualisation of complex needs[2, 9, 12]. However, the present study extends existing frameworks by emphasising the significance of trauma histories, prolonged episodes of rough sleeping, and repeated interactions with the criminal justice system as critical factors that compound the complexity of individuals’ needs.

This study revealed that PHECHS have experienced significant childhood trauma, the effects of which endure into adulthood. The most common types of childhood trauma identified among PHECHS include sexual abuse, physical abuse and being raised in environments where important adults abuse substances or are involved in criminal activities. These findings are consistent with the existing literature, including Mabhala et al [6] research, which systematically analysed and reported the relationships between adverse childhood experiences and later-life outcomes[6, 7, 21]. For instance, Mabhala et al identified clusters of traumatic exposures that serve as significant predictors of homelessness [6]. The principal scholarly contribution of the present investigation lies in its elucidation that such adversities exhibit the strongest correlation with chronic, entrenched homelessness.

Furthermore, the analysis demonstrates that individuals experiencing persistent and entrenched homelessness (PHECHS) are disproportionately affected by mental health disorders and frequently engage in maladaptive behaviours, such as substance use, as strategies to mitigate psychological distress and trauma. These behaviours not only serve as coping mechanisms but also contribute to the initial onset and perpetuation of homelessness. Prolonged exposure to homelessness significantly amplifies psychological morbidity, creating a self-reinforcing cycle that impedes recovery and reintegration. Consequently, participants emphasised the necessity of adopting trauma-informed and sustained, attritional interventions tailored to the complex needs of this population.

The analysis demonstrates that PHECHS experience structural disadvantages, notably persistent poverty, child poverty, limited educational attainment, chronic unemployment and deficits in social and personal skills. Collectively, these interconnected determinants not only critically impede access to health and social services and substantially increase the risk of housing insecurity but also deprive individuals of most fundamental human rights[16, 22–24].These include the right to social security, the right to food and housing as part of an adequate standard of living, labour rights, the right to health, and the right to education and participation in cultural life [16]. This necessitates a paradigm shift from viewing homelessness solely as a lack of accommodation requiring remedial housing interventions to recognising the need for structural reforms and a human rights-based approach.

Evidence indicates that sustained, structural-level interventions—underpinned by a coherent legislative framework and sufficient financial investment—are most effective in reducing homelessness. Between 1997 and 2010, the UK government undertook a coordinated structural response, most notably through the establishment of the Social Exclusion Unit (SEU) within the Prime Minister’s office, thereby elevating homelessness to a top policy priority[1]. The SEU’s primary mandate was to address the structural determinants of social exclusion, such as unemployment, low educational attainment, homelessness and rough sleeping, health inequalities, crime, and anti-social behaviour [1, 2]. These targeted efforts led to the lowest levels of homelessness ever recorded in the UK. By 2010, this achievement was reflected in official statistics, with 48,010 children residing in temporary accommodation [1, 3]

Participants strongly asserted that homelessness is rooted not only in the absence of shelter, but more fundamentally in deficits in essential practical skills necessary for securing and maintaining stable accommodation. They identified a range of skill gaps—including employment-seeking, financial management, navigating health and social services, self-management, time management, and adherence to healthcare appointments—that impede individuals’ ability to sustain housing. Recognising homelessness as a multifaceted phenomenon, rather than a simple lack of physical shelter, informed a series of integrated policy interventions implemented between 1998 and 2010[27–31]. This holistic approach proved highly effective, contributing to the near eradication of homelessness in the UK during that period. These insights emphasise the need for targeted, skill-based interventions as one of several important elements for promoting long-term housing stability and addressing the structural determinants of homelessness.

## 5. Limitations of the study

This study employed a qualitative methodology, analysing data through the lens of socioeconomic determinants of health. While this approach offers valuable insights, it also introduces inherent limitations. The reliance on a predetermined theoretical framework may have shaped both data collection and analysis, potentially narrowing the scope and objectivity of the findings. As a result, interpretations and conclusions are closely tied to the selected analytical perspective. Furthermore, our theoretical explanation constitutes an interpretation of participants’ accounts regarding the causes and circumstances of homelessness, rather than an objective or universally applicable explanation.

The study was conducted in an affluent, predominantly white British community in the Northwest of England. This demographic context is not fully representative of the broader population of England. Consequently, both the local context and the specific services available to individuals experiencing homelessness may have influenced participants’ perspectives and experiences, limiting the transferability of the findings to other settings.

Additionally, the study primarily focuses on individuals experiencing the most severe form of homelessness, a group that comprises only 4% of the overall homeless population. This focus may further limit the generalizability of the findings to the wider population of people experiencing homelessness.

Certain population groups were underrepresented in this study, notably individuals from ethnic minority backgrounds, women, and young people. This underrepresentation may have resulted in a limited range of perspectives and experiences, further constraining the comprehensiveness and generalizability of the findings.

## 6. Conclusion

This study demonstrates that homelessness among individuals with complex health and social needs arises from cumulative lifelong adversities, entrenched social and economic inequalities, and persistent systemic failures, rather than from isolated incidents. The analysis reveals that homelessness is not solely attributable to the lack of shelter, but is more fundamentally linked to deficits in essential practical skills required to secure and maintain stable accommodation. Specifically, the study identifies significant skill gaps—including employment-seeking, financial management, navigation of health and social services, self-management, time management, and adherence to healthcare appointments—that collectively impede individuals’ capacity to sustain housing.

The findings assert that addressing homelessness extends beyond the provision of emergency accommodation or fragmented interventions. Instead, an effective response requires a comprehensive, sustained, and rights-based strategy that addresses the structural determinants of homelessness, such as poverty, trauma, restricted access to care, and social exclusion. The evidence signifies the need for multiagency collaboration, trauma-informed practice, and a sustained commitment to human rights to achieve lasting systemic change. By framing homelessness as a violation of fundamental rights, the study locates the primary responsibility for redress with the state as the rights-bearer. Furthermore, it demonstrates that reductions in homelessness rates are consistently linked to the implementation of robust legislative frameworks, national guidelines, and substantial financial investment.

## 7. Policy points

- Homelessness undermines fundamental human rights, including the right to social security, adequate food and housing, fair labour conditions, health, education, and cultural participation.
- Therefore, the governments, as duty-bearers of human rights, are both morally and legally obligated to prevent and eradicate homelessness.
- An effective response demands a comprehensive, sustained, and rights-based strategy that tackles the root causes of homelessness, including poverty, trauma, and social exclusion.
- Homelessness is fundamentally linked to deficits in essential life skills needed to obtain and maintain stable housing. Key skill gaps include job seeking, financial management, navigating health and social services, self-management, time management, and keeping healthcare appointments, all of which hinder individuals’ ability to sustain housing.
- Enduring reductions in homelessness are achieved through robust legislation, clear national guidelines, and significant financial investment.

## Data Availability

All data produced in the present study are available upon reasonable request to the authors

## References

1. HH Group. *Changing faces of homelessness in the UK: 2025 trends*. 2025 [cited 2025 11/11/2025]; Available from: https://homelesshostelstaff.co.uk/uk-homelessness/#:∼:text=You%20might%20be%20wondering%20what%E2%80%99s%20happening%20with%20homelessness,working%2C%20and%20the%20overall%20impact%20on%20people%E2%8 0%99s%20lives.

2. Rosengard, A., et al., A literature review on multiple and complex needs. Scottish Executive Social Research, 2007.

3. Watson, L. and P. Edelman, Improving the juvenile justice system for girls: Lessons from research and practice. Georgetown Journal on Poverty Law & Policy, 2012. 19: p. 521–530.

4. McCann, D., R. Bull, and T. Winzenberg, The daily patterns of time use for parents of children with complex needs:A systematic review. Journal of Child Health Care, 2012. 16(1): p. 26–52.

5. Macías Balda, M., Complex needs or simplistic approaches? Homelessness services and people with complex needs in Edinburgh. Social Inclusion, 2016. 4(4): p. 28–38.

6. Mabhala, M., et al., Homelessness Is Socially Created: Cluster Analysis of Social Determinants of Homelessness (SODH) in North West England in 2020. Int J Environ Res Public Health, 2021. 18(6).

7. Mabhala, M.A., A. Yohannes, and M. Griffith, Social conditions of becoming homelessness: qualitative analysis of life stories of homeless peoples. International Journal for Equity in Health, 2017. 16(1): p. 150.

8. Harland, J.M., et al., Understanding the life experiences of people with multiple complex needs: peer research in a health needs assessment. European Journal of Public Health, 2022. 32(2): p. 176–190.

9. Keene, J., Complex needs: interprofessional pracrice. 2001, London: Wiley Publication.

10. Rankin, J. and S. Regan, *Meeting complex needs in social care.* Housing, Care and Support, 2004. 7(3): p. 4–8.

11. Padgett, D.K., et al., Complex recovery: Understanding the lives of formerly homeless adults with complex needs. Journal of Social Distress and Homelessness, 2016. 25(2): p. 60–70.

12. Macías Balda, M., Complex Needs or Simplistic Approaches? Homelessness Services and People with Complex Needs in Edinburgh. Social Inclusion, 2016. 4(4): p. 28–38.

13. 13. National Institute for Health and Care Excellence. Social work with adults experiencing complex needs. 2022 [cited 2026 20/02/2026]; Available from: https://www.nice.org.uk/guidance/ng216/resources/social-work-with-adults-experiencing-complex-needs-pdf-66143778560197.

14. Health and Care. What are complex needs*?* 2025 [cited 2025 22/10/2025]; Available from: https://www.oneadvanced.com/resources/what-are-complex-needs/.

15. Care for Family. Complex Needs: What is it? Examples & Care Information. 2025; Available from: https://info.careforfamily.com.au/blog/complex-needs-what-is-it-examples.

16. International Covenant on Economic Social and Cultural Rights. International Covenant on Economic, Social and Cultural Rights Adopted and opened for signature, ratification and accession by General Assembly resolution 2200A (XXI) of 16 December 1966 entry into force 3 January 1976, in accordance with article 27 1966; Available from: https://www.ohchr.org/en/instruments-mechanisms/instruments/international-covenant-economic-social-and-cultural-rights.

17. Horrigan-Kelly, M., M. Millar, and M. Dowling, Understanding the Key Tenets of Heidegger’s Philosophy for Interpretive Phenomenological Research. International Journal of Qualitative Methods, 2016. 15(1): p. 1609406916680634.

18. Larkin, M. and A.R. Thompson, Interpretative Phenomenological Analysis in Mental Health and Psychotherapy Research. Qualitative Research Methods in Mental Health and Psychotherapy, 2011. 13(47): p. 1–8.

19. Willig, C. and A. Billin, Existentialist-Informed Hermeneutic Phenomenology, in Qualitative Research Methods in Mental Health and Psychotherapy. 2011. p. 117–130.

20. Benner, P., Interpretive Phenomenology: Embodiment, Caring, and Ethics in Health and Illness. 1994, London: SAGE Publications, Inc.

21. Mabhala, M.A. and A. Yohannes, Being at the Bottom Rung of the Ladder in an Unequal Society: A Qualitative Analysis of Stories of People without a Home. International journal of environmental research and public health, 2019. 16(23): p. 4620.

22. United Nations. Homelessness and human rights - Special Rapporteur on the right to adequate housing. 2024 [cited 2024 16/07/2024]; Available from: https://www.ohchr.org/en/special-procedures/sr-housing/homelessness-and-human-rights.

23. United Nations. Universal Declaration of Human Rights 1948 [cited 2025 2710/2025]; Available from: https://www.un.org/en/about-us/universal-declaration-of-human-rights.

24. Office of the United Nations High Commissioner for Human Rights. Convention on the Rights of the Child. 1989 [cited 2025 28/10/2025]; Available from: https://www.ohchr.org/sites/default/files/Documents/ProfessionalInterest/crc.pdf.

25. Legislation.gov.uk, *Housing Act of* 1996. 1996, Uk Government: London.

26. Shelter. Human rights challenges to local authority homelessness decisions. 2021 [cited 2026 17/02/2026]; Available from: https://england.shelter.org.uk/professional_resources/legal/homelessness_applications/homelessness_reviews_and_appeals/human_rights_challenges_to_local_authority_homelessness_decisions#source-5.

27. Christie, D.A. and N.J. Crowson. New Labour and Street Homelessness 1997-2010. 2025 [cited 2025 14/11/2025]; Available from: https://historyandpolicy.org/policy-papers/papers/new-labour-and-street-homelessness-1997-2010/.

28. Atkinson, A. and J. Hill. *Exclusion, Employment and Opportunity*. 1998 [cited 2025 18/11/2025]; Available from: https://sticerd.lse.ac.uk/dps/case/cp/Paper4.PDF.

29. Kennedy, S. Social exclusion: Social Policy Section 2007 [cited 2025 18/11/2025]; Available from: https://researchbriefings.files.parliament.uk/documents/SN04221/SN04221.pdf.

30. Communities and Local Government. Statutory Homelessness: March Quarter 2010 *England*. 2010 [cited 2025 18/1/2025]; Available from: https://assets.publishing.service.gov.uk/media/5a74f089ed915d502d6cc391/1611050.pdf?utm_sour ce=chatgpt.com.

31. National Audit Office. *More than a roof:vProgress in tackling homelessness*. 2005; Available from: https://webarchive.nationalarchives.gov.uk/ukgwa/20170207052351/ https://www.nao.org.uk/wp-content/uploads/2005/02/0405286es.pdf.

